# Clinical Validation of a Novel T-cell Receptor Sequencing Assay for Identification of Recent or Prior SARS-CoV-2 Infection

**DOI:** 10.1101/2021.01.06.21249345

**Authors:** Sudeb C. Dalai, Jennifer N. Dines, Thomas M. Snyder, Rachel M. Gittelman, Tera Eerkes, Pashmi Vaney, Sally Howard, Kipp Akers, Lynell Skewis, Anthony Monteforte, Pam Witte, Cristina Wolf, Hans Nesse, Megan Herndon, Jia Qadeer, Sarah Duffy, Emily Svejnoha, Caroline Taromino, Ian M. Kaplan, John Alsobrook, Thomas Manley, Lance Baldo

## Abstract

**Background:** While diagnostic, therapeutic, and vaccine development in the COVID-19 pandemic has proceeded at unprecedented speed and scale, critical gaps remain in our understanding of the immune response to SARS-CoV-2. Current diagnostic strategies, including serology, have numerous limitations in addressing these gaps. Here we describe clinical performance of T- Detect™ COVID, the first reported assay to determine recent or prior SARS-CoV-2 infection based on T-cell receptor (TCR) sequencing and immune repertoire profiling from whole blood samples.

**Methods:** Methods for high-throughput immunosequencing of the TCRβ gene from blood specimens have been described^1^. We developed a statistical classifier showing high specificity for identifying prior SARS-CoV-2 infection^2^, utilizing >4,000 SARS-CoV-2-associated TCR sequences from 784 cases and 2,447 controls across 5 independent cohorts. The T-Detect COVID Assay comprises immunosequencing and classifier application to yield a qualitative positive or negative result. Several retrospective and prospective cohorts were enrolled to assess assay performance including primary and secondary Positive Percent Agreement (PPA; N=205, N=77); primary and secondary Negative Percent Agreement (NPA; N=87, N=79); PPA compared to serology (N=55); and pathogen cross-reactivity (N=38).

**Results:** T-Detect COVID demonstrated high PPA in subjects with prior PCR-confirmed SARS-CoV-2 infection (97.1% 15+ days from diagnosis; 94.5% 15+ days from symptom onset), high NPA (∼100%) in presumed or confirmed SARS-CoV-2 negative cases, equivalent or higher PPA than two commercial EUA serology tests, and no evidence of pathogen cross-reactivity.

**Conclusion:** T-Detect COVID is a novel T-cell immunosequencing assay demonstrating high clinical performance to identify recent or prior SARS-CoV-2 infection from standard blood samples. This assay can provide critical insights on the SARS-CoV-2 immune response, with potential implications for clinical management, risk stratification, surveillance, assessing protective immunity, and understanding long-term sequelae.

## INTRODUCTION

The emergence and rapid spread of SARS-CoV-2, the virus that causes coronavirus disease (COVID-19), has resulted in a global pandemic of over 75 million cases and 1.7 million deaths worldwide in 2020^3^. Despite rapidly accumulating data and recent approvals of vaccines, key gaps remain in our understanding of the immune response to SARS-CoV-2, including the nature and durability of the correlates of protection, the relationship between immune response and individual disease susceptibility and severity, and the possibility that some immune phenotypes may be more advantageous or efficient at preventing infection or severe disease^4–6^.

Such knowledge gaps translate into critical areas of unmet need in the diagnosis and management of COVID-19 and epidemiologic monitoring of the pandemic. Currently, serologic (antibody) testing of IgM, IgG, and/or IgA isotypes is the primary modality for evaluating prior SARS-CoV-2 infection or exposure, disease prevalence and incidence, and immune protection^7–9^. While antibody testing has been shown to capture a larger percentage of exposures than PCR testing^10^, its performance has a number of limitations, including low or absent antibody titers in individuals with asymptomatic or mild infection^11,12^, declines in antibody levels over time^13,14^, and false-positive results from cross-reactivity to other viruses, infections, or unrelated autoimmune conditions^15–17^. There is also wide variability in performance across the numerous SARS-CoV-2 serologic tests currently available^18^. In addition, it remains unclear whether the results of antibody testing correlate with long-term protective immunity or prevention of transmission^9^. Finally, serologic testing may not reflect the true extent of individual pre-existing immunity, as SARS-CoV-2-reactive T-cells have been identified in 20-50% of individuals with no known exposure^19–21^. These issues have severely limited the utility of serologic testing to inform individual risk or guidance of public behavior, including physical distancing, mask wearing or resumption of activities^22^.

Recent reports showing declining levels of anti-SARS-CoV-2 IgG and neutralizing antibodies only a few months after infection, particularly in asymptomatic individuals, have fueled concerns that achieving long-term immunity to SARS-CoV-2, whether by natural infection or by vaccination, will be challenging^11,13,14^. This is supported by accumulating data regarding SARS-CoV-2 reinfections in as few as 2 months following initial infection^23,24^. The observation that pan-Ig antibody titers appear stable up to 4 months following diagnosis suggests that long-term immunity to SARS-CoV-2 involves complex and multifactorial mechanisms, including the action of long-lived plasma cells and coordination between the humoral and cellular immune responses^7,10^. It is not known what proportion of exposed individuals will exhibit a memory antibody response, although early data suggest that ∼10% of individuals recover from SARS-CoV-2 infection yet have no detectable antibodies^25,26^.

In addition to the humoral response, cellular responses play a central role in SARS-CoV-2 immunity^10,27^. Indeed, the majority of patients diagnosed with COVID-19, including convalescent patients across a wide spectrum of disease severity, generate CD8+ and CD4+ T-cell responses^19,28^, which have been associated with milder disease and protection from infection^29,30^. T cells also play a critical role in activating the humoral response and can precede antibodies to serve as the first sign of the immune response to SARS-CoV-2 infection, particularly in asymptomatic or mild illness^20,31^. SARS-CoV-2–specific T cells are persistent, remaining elevated at least 6 months post-infection, in some cases in the absence of seroconversion^2,32–34^. Finally, aberrant T- or B-cell responses have been implicated in the immune dysregulation underlying severe COVID and inflammatory sequelae including multisystem inflammatory syndrome in children (MIS-C) and in adults (MIS-A)^35,36^, myocardial involvement^37,38^, and post-acute syndromes such as “long COVID^39^.” Exploration of the precise contributions and timing of both humoral and cellular responses is needed to fully understand the biological basis for long-term immunity to SARS-CoV-2 infection and its associated complications. This has become particularly salient with the advent of mass vaccination strategies for COVID-19, where identifying the correlates of vaccine-mediated immunity is central to assessing the durability of protection, whether pre-existing immunity influences the vaccine response^40^, and whether recently-reported viral escape mutants with enhanced infectivity^41^ can evade vaccine-induced immunity.

A number of features inherent to the biology of the T-cell immune response make it a desirable target for identifying and tracking disease exposure. The cellular immune response is: 1) sensitive to very small amounts of antigen; 2) specific, binding only to specific antigens; 3) naturally amplified through clonal expansion; 4) systemic, as T-cell clones circulate in the blood; and 5) persistent, as it is maintained in long-term memory. Here we describe the implementation and extensive clinical validation of T-Detect™ COVID, a novel high-throughput assay to determine recent or prior SARS-CoV-2 infection based on T-cell receptor gene sequencing and subsequent repertoire profiling from whole blood samples, following US Food and Drug Administration guidance “*Policy for Coronavirus Disease-2019 Tests During the Public Health Emergency (Revised) May2020.”* We demonstrate high positive and negative percent agreement of this assay to identify or exclude prior SARS-CoV-2 infection in PCR-confirmed SARS-CoV-2 cases across several cohorts and longitudinal timepoints. We also show that the assay has equivalent or better performance than commercially-available EUA antibody tests at all timepoints evaluated^42^, and lacks cross-reactivity to several viral and/or respiratory pathogens.

## METHODS

### Ethics

All samples were collected pursuant to an Institutional Review Board (IRB)-approved clinical study protocol. For residual samples collected under prospective study protocols, informed consent was obtained from participants. All other samples from cohorts described below were collected as clinical remnant samples. (See Supplement for detailed information).

### Clinical Cohorts

Clinical specimens were collected via distinct study arms: 1) a retrospective arm with SARS-CoV-2 positive and negative residual samples from prior research studies and remnant clinical samples; and 2) a prospective arm to collect samples from participants with symptoms compatible with COVID-19 and testing either positive or negative by SARS-CoV-2 RT-PCR. These two study arms provided samples to demonstrate the clinical agreement of the T-Detect™ COVID Assay to determine the PPA and NPA. Study populations are described below and in the Supplement.

#### PPA Study Cohorts

The primary PPA study evaluated residual blood samples (N=222) from subjects diagnosed with SARS-CoV-2 infection based on the EUA Abbott RealTime SARS-CoV-2 RT-PCR test from a single US reference lab (New York) (Table 1).

**Table 1.** Description of RT-PCR positive SARS-CoV-2 samples used for primary & secondary analyses.

|  | **Primary Analyses** | **Secondary Analyses** |  |
| --- | --- | --- | --- |
| **Cohort name** | Discovery Life Sciences (DLS)* | ImmuneRACE* | ImmuneSense COVID-19* |
| **Cohort information** | Clinical remnant samples from subjects that were positive for SARS-CoV-2 | Retrospective use of residual samples from a prior research study with confirmed SARS-CoV-2 infection via medical record search (NCT04494893) | Prospective collection of individuals being tested for SARS-CoV-2, included participants that tested positive for SARS-CoV-2 (NCT04583982) |
| **Number of unique samples** | N = 222 | N = 69 | N = 8 |
| **Study population** | Basic demographics, from a New York reference lab | Enrolled ages 18-89, samples collected nationwide, 24 virtual locations throughout the US | Enrolled ages 18-89, two clinical drive-thru testing sites in New Jersey |
| **Sample types**<br>**PCR Comparator**<br>**test** | Frozen whole blood<br>Abbott RT-PCR<br>SARS-CoV-2 EUA | Frozen whole blood<br>Multiple independent<br>EUA authorized test<br>methods | Frozen whole blood<br>Abbott RT-PCR<br>SARS-CoV-2 EUA |

\*A detailed description of these cohorts is provided in the Supplement

Secondary PPA assessments were performed using both retrospectively and prospectively collected samples from multiple cohorts (N=77; ImmuneRACE and ImmuneSense™ COVID-19 cohorts, Supplement) and identified as positive based on a variety of EUA testing methods performed by a number of different labs. Given the potential for variability in RT-PCR performance given the use of numerous tests by multiple labs, samples were categorized by days since symptom onset (Table 1).

#### NPA Study Cohorts

The primary NPA included 124 retrospective frozen clinical remnant blood samples collected prior to December 2019 (Table 2) and thus presumed negative for SARS-CoV-2 infection. These samples were collected over two years, during all months (including cold/flu season), and from diverse geographical areas in the United States (Table 2).

**Table 2.** Description of SARS-CoV-2 negative samples for primary and secondary NPA.

|  | **Primary NPA** | **Secondary NPA** |
| --- | --- | --- |
| **Cohort Name** | Discovery Life Sciences (DLS)* | ImmuneSense COVID-19* |
| **Cohort Details** | Retrospective collection | Prospective collection |
| **Number of Unique Negative Samples** | N=124 | N=79 |
| **Study population** | Diverse populations collected within the US upon presentation to clinic with a variety of symptoms, including respiratory illnesses | Single site collection, New Jersey |
| **Dates of collection** | Jul. 2017 – Nov. 2019 | Oct. - Dec. 2020 |
| **Sample types** | Frozen blood | Frozen blood |
| **Nasopharyngeal Test** |  | Abbott RT-PCR SARS-CoV-2 EUA |
| **Comparators test at time of collection** |  | BioFire RP 2.1 EUA |
| **Antibody Test Comparators at time of collection** |  | Abbott Architect SARS-CoV-2 IgG<br>Roche Elecsys Anti-SARS-CoV-2 |

\*A detailed description of these cohorts is provided in the Supplement

The secondary NPA study included blood samples from subjects enrolled prospectively (ImmuneSense COVID-19) from Oct-Nov 2020 who presented with SARS-CoV-2 symptoms but tested negative for SARS-CoV-2 using RT-PCR EUA, BioFire RP V2.1, and EUA antibody tests (Table 2).

### Clinical Specimens

From all sources, whole blood samples were collected in EDTA tubes, frozen, and shipped to Adaptive for immunosequencing. Paired serum samples were tested using two different EUA antibody assays: 1) Elecsys® Anti-SARS-CoV-2; Roche (all isotypes); and 2) SARS-CoV-2 Antibody, IgG; LabCorp. Detailed serology assay information is in the Supplement.

### Classifier Development and Training

We have previously described the development of a SARS-CoV-2 classifier based on TCRβ DNA sequences from blood samples^2^. Briefly, one-tailed Fisher’s exact tests were performed on all unique TCR sequences comparing their presence in SARS-CoV-2 PCR-positive samples (n=784) with negative controls (n=2,447) to generate a list of SARS-CoV-2-associated sequences which are exclusive to, or greatly enriched, in PCR-positive samples. These sequences were used to create a classifier by logistic regression with two dependent variables, the number of unique TCRβ DNA sequences encoding a SARS-CoV-2-associated sequence and the total number of unique TCRβ DNA sequences in the sample. The diagnostic model threshold is set to demonstrate 99.8% specificity against a set of 1,657 held out negative controls not used in training^2^.

### T-Detect COVID Assay

#### Process Overview

The T-Detect COVID Assay consists of 1) a core assay designed to sequence and quantify rearranged TCRβ sequences from gDNA extracted from peripheral blood and 2) diagnostic software, which applies a COVID-specific algorithm to the TCRβ sequence repertoire data to determine a result. The system consists of reagents, instrumentation, software and instructions needed to perform the process steps as summarized in Figure 1.

**Figure 1.**
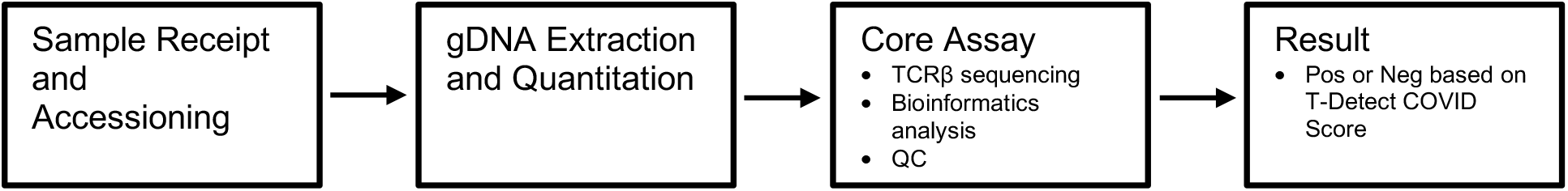
T-Detect COVID Assay process overview.

#### Sample Collection and Processing

Peripheral whole blood is collected in a 10mL EDTA vacutainer tube and shipped overnight at ambient temperature to the Adaptive clinical laboratory. Upon receipt it is accessioned and stored refrigerated at 4C until processed that same day via automated gDNA extraction or stored frozen at –80C if extraction is at a later date.

#### Sample and Library Preparation, Sequencing, and Pipeline Analysis

Detailed methods for sample preparation, immunosequencing, and pipeline analysis have been described previously^1,2^. Briefly, a target gDNA sample input of 18µgs is isolated from 2mL of fresh or frozen peripheral whole blood (6mL is requested). This target gDNA input ensures that samples achieve a minimum unique productive rearrangements (UPR) input QC specification. A multiplex PCR strategy with synthetic TCRβ molecules added to each reaction is used to amplify rearranged TCRβ sequences from gDNA. PCR libraries are loaded together on a single sequencing run and sequencing performed using the Illumina NextSeq 500/550 System. Sequence data are extracted and reads are attributed to data derived from biological vs. synthetic templates to derive template estimates for each identified receptor sequence as well as input cell counts.

#### T-Detect COVID algorithm

The COVID-specific algorithm (classifier) which was developed as described above and locked prior to initiating any of the T-Detect COVID validation studies is applied to the core assay output. The classifier identifies and quantifies any SARS-CoV-2-associated TCRs from a predetermined list of several thousand SARS-CoV-2-associated TCRs and also quantifies non-SARS-CoV-2 TCR sequences. These factors are mathematically combined into a score representing the relative enrichment for SARS-CoV-2-associated TCR sequences. This score is compared to a pre-specified threshold derived during algorithm training to classify the patient sample as positive or negative for an immune response to SARS-CoV-2.

## RESULTS

### Public enhanced sequences associated with SARS-CoV-2 infection distinguish cases from controls

Initial development of the COVID classifier utilized public enhanced SARS-CoV-2 sequences from two cohorts, Discovery Life Science (DLS, from New York, USA) and NIH/NIAID (from Italy), comprising a total of 483 cases, with 1,798 controls collected before the emergence of SARS-CoV-2 in 2020. A total of 1,828 enhanced SARS-CoV-2 sequences were identified from this first dataset which collectively distinguish cases from controls (Figure 2a). Notably, these enhanced sequences were also substantially enriched in 397 cases from three additional held-out cohorts: ISB (Institute of Systems Biology’s Covid-19 Immune Response Study; Seattle, WA), H12O = (Hospital 12 de Octubre; Madrid, Spain), and BWNW = (Bloodworks Northwest; Seattle, WA) but not seen at the same elevated rates in 1,702 additional held-out controls (Figure 2b).

**Figure 2.**
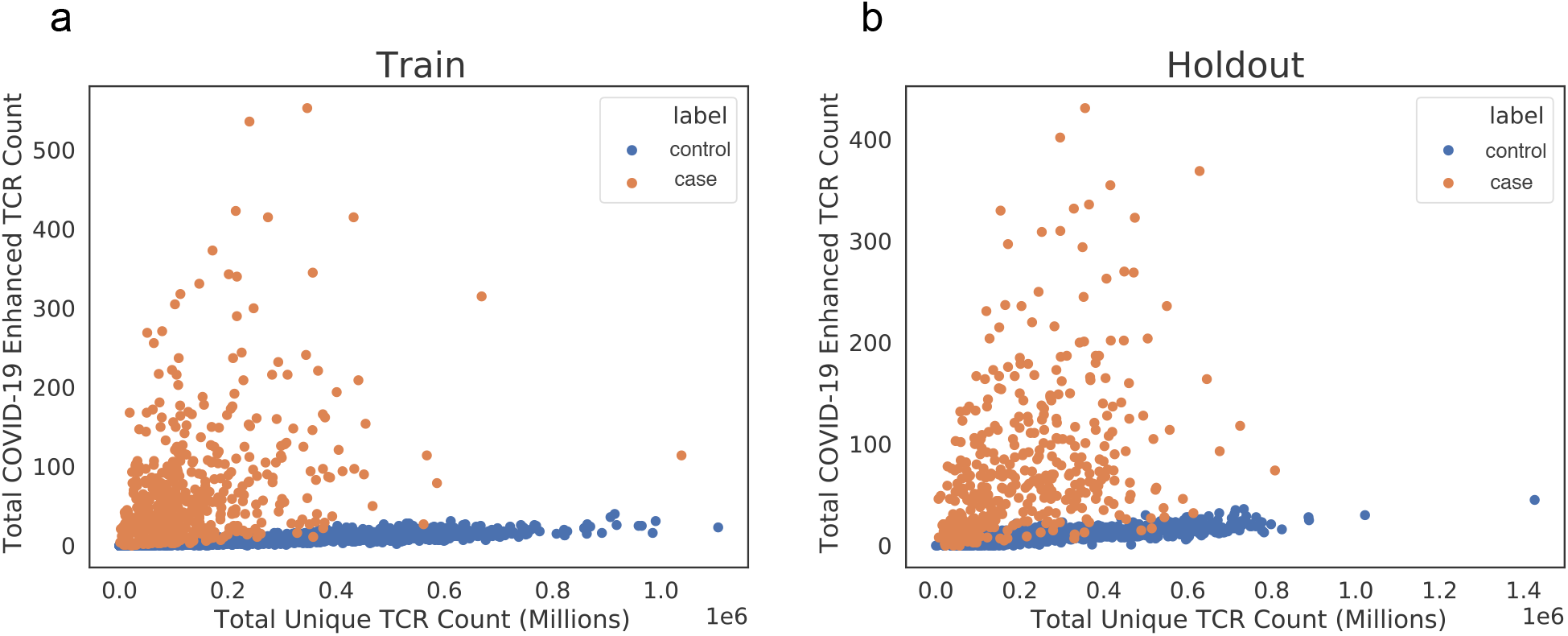
Public enhanced sequences associated with SARS-CoV-2 infection distinguish cases from controls. Panels (a) and (b) show the number of TCRβ DNA sequences in a subject that encode a SARS-CoV-2 enhanced sequence versus the total number of unique TCRβ DNA sequences sampled from that subject for a large number of cases and controls. Panel (a) represents the training set to identify this initial list of enhanced sequences (DLS and NIH/NIAID cohorts), and panel (b) represents a hold-out set with no overlap with the training set (ISB, H12O and BWNW cohorts). Both panels show a similar number and separation of enhanced sequences in cases versus controls.

As additional data enabled identification of more SARS-CoV-2 associated TCRs to improve performance of the classifier^2^, our final classifier was trained using 784 cases from all five cohorts referenced above (and in Supplemental Table 1), as well as 2,447 controls. We then set the diagnostic model threshold to 99.8% specificity on an independent set of 1,657 negative controls not used in training. The final classifier includes a total of 4,470 SARS-CoV-2 associated sequences. The classifier’s performance appears robust to potential confounders such as age and sex (Figure 3a,b), and its performance has been tested in several independent studies^2,42^, suggesting equal or better sensitivity to antibody serology testing.

**Figure 3.**
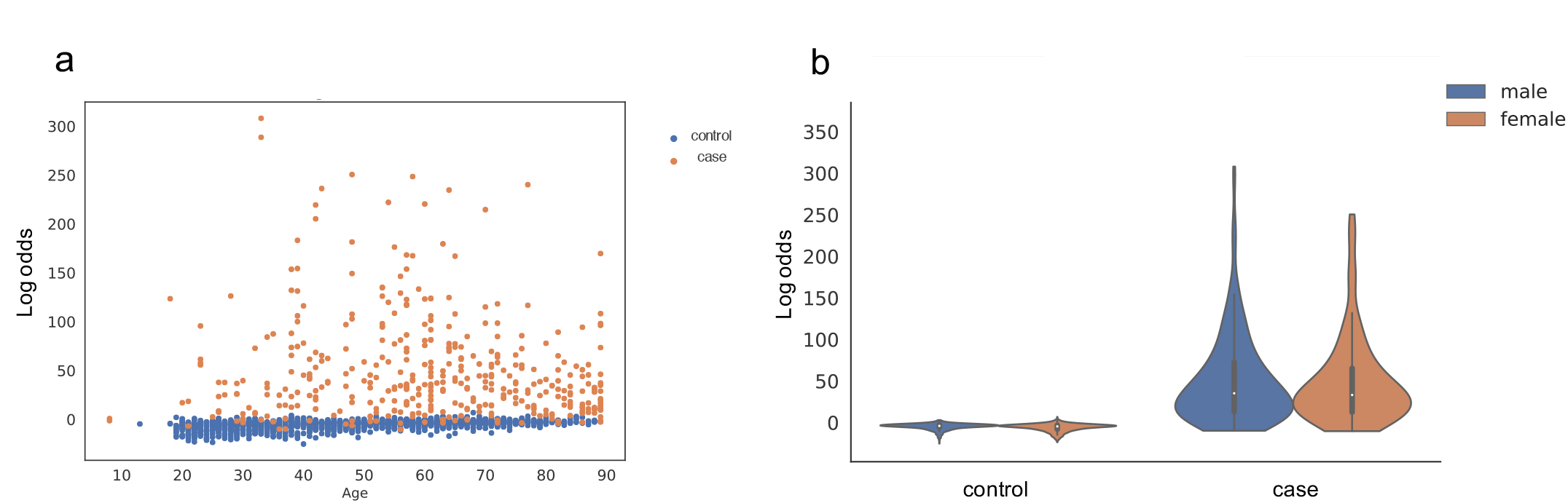
Performance of T-cell classifier to separate SARS-CoV-2 cases from controls is consistent across age and gender. Performance of T-cell classifier to separate SARS-CoV-2 cases from controls is consistent across ages (a) and in both males and females (b). Both plots report model scores as the untransformed log-odds estimated from the logistic regression classifier. The violin plot in panel (b) visualizes the density of log-odds scores among male and female cases and controls, with median and interquartile range values indicated.

### High Positive Percent Agreement (PPA) with SARS-CoV-2 PCR

Two separate positive percent agreement (PPA) studies were undertaken to evaluate T-Detect COVID Assay performance in subjects with confirmed positive SARS-CoV-2 PCR: a primary PPA analysis relative to days since diagnosis and a secondary PPA analysis relative to days from symptom onset. In the primary PPA study, 205/222 samples tested were from unique subjects and passed all QC and threshold requirements making them eligible for analysis. In the secondary PPA study, all 77 samples tested were from unique individuals, passed QC and threshold requirements, and were included for analysis. Samples were tested out to a maximum of 106 days from symptom onset. The PPA for various timepoints is displayed in Table 3. PPA for the T-Detect COVID Assay was highest (97.1%) in the timeframe of ≥15 days since diagnosis as well as ≥15 days since symptom onset (94.5%). (Table 3).

**Table 3.** Positive Percent Agreement (PPA) of T-Detect COVID Assay with SARS-CoV-2 RT-PCR according to days since symptom onset or days since diagnosis.

| **Days Since**<br>**Diagnosis** | **RT-PCR+**<br>**Samples (N)** | **T-Detect Positive (N)** | **T-Detect PPA (95% CI)** |
| --- | --- | --- | --- |
| 0-7 days | 35 | 25 | 71.4 (53.7 - 85.4) |
| 8-14 days | 33 | 31 | 93.9 (92.7 - 99.3) |
| ≥15 days | 137 | 133 | 97.1 (92.7 - 99.2) |
| All (range 0-91 days) | 205 | N/A | N/A |
| **Days Since**<br>**Symptom Onset** |  |  |  |
| 0-7 days | 13 | 7 | 53.8 (25.1 - 80.8) |
| 8-14 days | 9 | 7 | 77.8 (40.0 - 97.2) |
| ≥15 days | 55 | 52 | 94.5 (84.9 - 98.9) |
| All (range 0-106 days) | 77 | N/A | N/A |

### High Negative Percent Agreement (NPA) in presumed and/or confirmed SARS-CoV-2 negative samples

Two separate negative percent agreement (NPA) studies were undertaken to evaluate T-Detect COVID Assay performance: a primary NPA analysis of retrospectively sourced whole blood samples from pre-pandemic timepoints (July 2017- Nov 2019) and thus presumed SARS-CoV-2 negative, and a secondary NPA analysis of prospectively collected samples from symptomatic but SARS-CoV-2 test negative subjects. In the primary NPA study, 87 of 124 samples were from unique individuals, passed all standard QC and assay threshold requirements, and were used for analysis, yielding an NPA of 100% (Table 4). The majority of failure samples were due to failure to meet assay QC metrics or assay specific thresholds. Due to the retrospective sourcing of these samples, the collection conditions and biological/disease context of these samples was variable.

**Table 4.** Negative Percent Agreement (NPA) of T-Detect COVID Assay with pre-pandemic samples sourced retrospectively (DLS) and prospectively enrolled subjects (ImmuneSense COVID-19) negative for SARS-CoV-2 by EUA RT-PCR and antibody testing.

| Cohort | Samples (N) | T-Detect Negative<br>Results (N) | NPA (95% CI) |
| --- | --- | --- | --- |
| DLS | 87 | 87 | 100 (95.8 – 100) |
| ImmuneSense COVID-19 | 79 | 78 | 98.7 (93.1 – 99.97) |

The secondary NPA study assessed T-Detect COVID Assay performance prospectively in subjects presenting with compatible symptoms but testing negative for SARS-CoV-2 using RT-PCR (BioFire RP2.1 EUA) and EUA antibody tests. Of 79 subjects meeting these criteria, no samples failed QC or performance thresholds and all were included for analysis, yielding an NPA of 98.7% (Table 4).

### Equivalent or Greater PPA Than EUA Antibody Tests in Confirmed SARS-CoV-2 Cases

Additional analyses compared the PPA of T-Detect COVID Assay relative to results from serology-based antibody testing in paired SARS-CoV-2 positive samples from 77 unique subjects (EUA RT-PCR), and demonstrated PPA as high or higher than serology, particularly in early phases of infection (Table 5).

**Table 5.**
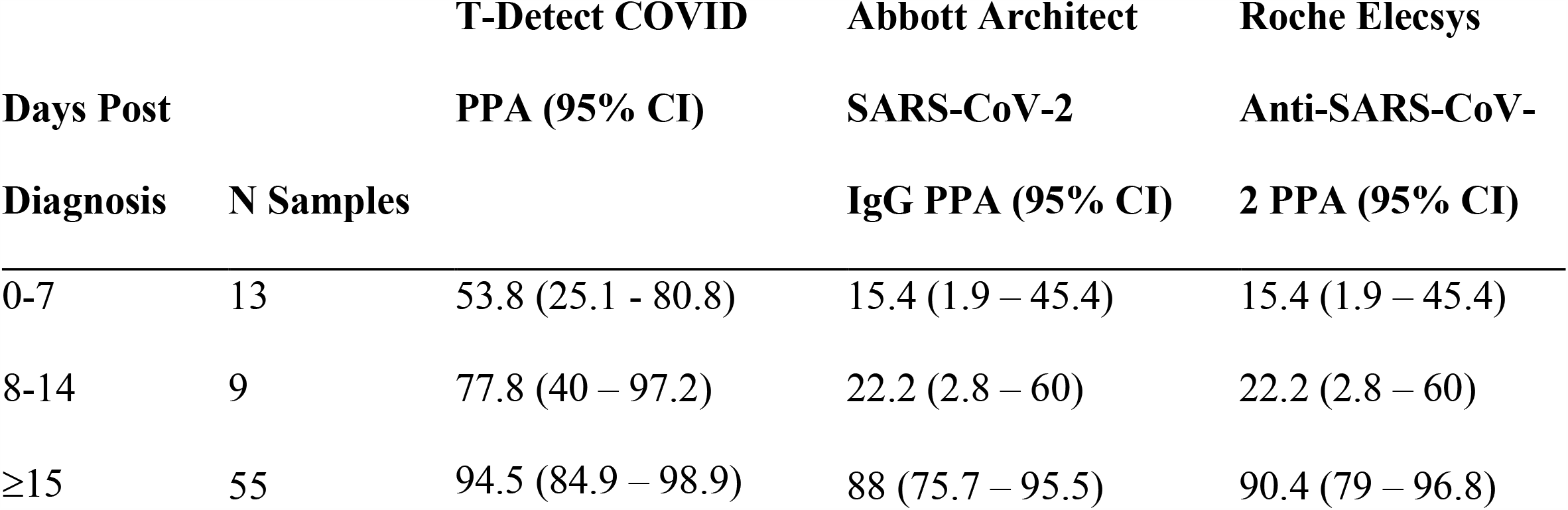
PPA of T-Detect COVID Assay results compared to serology-based assays in paired samples.

### Lack of cross-reactivity with other viruses/pathogens

The biology of the T-cell mediated response to infection inherently requires specificity between the TCRs in SARS-CoV-2 positive patient samples and the cognate antigens unique to SARS-CoV-2. The classifier development for this assay leveraged this biologic mechanism. The clinical call threshold was established by utilizing 1,657 controls/known negative samples collected in the U.S. prior to December 2019, from populations with a high prevalence of vaccination against, or infection with, potentially cross-reactive viruses. This approach yielded a clinical call threshold with an expected specificity of 99.8%.

Specificity was verified in a set of blood and PBMC samples collected from individuals infected with Influenza A/B, *Haemophilus influenzae b*, HIV, HBV and/or HCV to assess potential cross-reactivity. No samples tested positive using the T-Detect COVID Assay (Table 6).

**Table 6.**
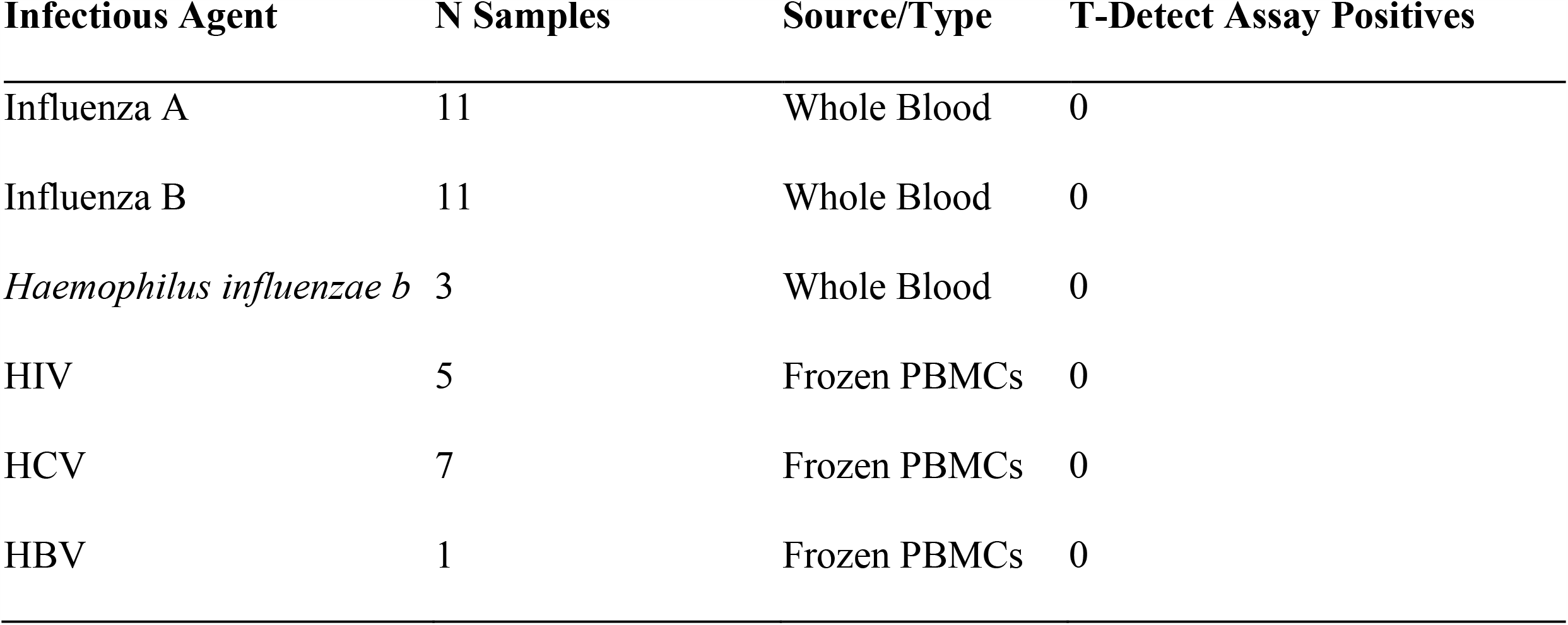
T-Detect COVID Assay results indicating 100% specificity (lack of cross reactivity) in individuals infected with Influenza A/B, *H. influenza b*, HIV, HCV and/or HBV.

## DISCUSSION

The COVID-19 pandemic has accelerated the development of myriad diagnostic testing strategies and platforms. Despite the critical roles of both humoral and cellular immune responses in SARS-CoV-2 infection and recovery, serologic testing is the predominant means of assessing previous infection, population-level prevalence and incidence, and potential immunity. Serology tests offer advantages of relatively low cost, fast turnaround time, and scalability; at the time of this publication, over 100 SARS-CoV-2 serologic tests are available for clinical use including over 60 with EUA status^43^. However, the limitations of serologic testing, including high variability in test performance across platforms and antibody isotypes tested^18^, waning or loss of antibody signal over time^11,13,14^, and absence of detectable antibodies in up to 10% of individuals including those with immunocompromising conditions^25,26^, expose unmet clinical and public health needs for immunologic testing strategies for SARS-CoV-2 that are consistent, durable, and more informative.

Using TCR gene sequencing from whole blood samples, we describe a sequence-based assay to identify recent or prior SARS-CoV-2 infection which demonstrates high PPA (>97% beyond 15 days following diagnosis), high NPA in presumed or confirmed negative SARS-CoV-2 infection (∼100%), equivalent or higher PPA compared to commercially available EUA serology tests, and lack of cross reactivity with a number of viral and/or respiratory tract pathogens. This performance was consistent across several retrospective and prospective cohorts and longitudinal sampling timeframes. Utilizing this approach in a real-world setting, we have shown previously that robust T-cell signals are persistent at least 6 months after primary SARS-CoV-2 infection^42^, consistent with other reports^44^. In the SARS-CoV-1 pandemic, detectable virus-specific T-cell responses were observed in recovered individuals up to 17 years later^21^. In direct real-world comparisons with serologic testing, we have observed up to a 20% lower sensitivity of commercially available antibody tests in identifying prior SARS-CoV-2 infection compared to T-Detect COVID, with greater reductions in serology performance occurring at later timepoints following infection^42^. Finally, we have reported a direct correlation between the magnitude of the measured SARS-CoV-2 T-cell response (in depth and breadth) and prior disease severity^11,42^.

These observations support the potential clinical utility of T-cell profiling in the COVID-19 pandemic as a means of risk stratification of disease progression and outcomes, detection of remote prior infection, informing public health and surveillance strategies, and clarifying the correlates of immune protection by providing a more comprehensive characterization of the immune response. We have previously applied our statistical classification framework based on immunosequencing data and T-cell repertoire profiling in determining CMV serostatus as a proof of principle^1^. The generation, validation, and application of different algorithms to immunosequencing data has the potential to yield clinical insights across multiple disease areas, particularly in infectious diseases and autoimmunity.

Robust T-cell profiling can also inform vaccine development. Vaccines targeting SARS-CoV-2 are capable of inducing type 1 helper T-cell (Th1) responses, in addition to high levels of binding and neutralizing antibodies that decline over time^4,45–47^. Indeed, Th1-skewed responses have been shown to drive protective humoral and T-cell responses in patients receiving vaccines directed against other viruses^48^. Thus, a combination of serological testing and high-throughput T-cell repertoire profiling could be beneficial for fully characterizing the nature of the immune response to SARS-CoV-2 vaccination, including assessment of the T-cell response and potential immune escape in recently-described viral variants that have evidence for increased infectivity and transmission^41^.

Finally, understanding the immune response to SARS-CoV-2 is critical for elucidating the etiology of immune dysregulation in severe COVID-19 and inflammatory sequelae. Recent data suggest that patients with severe COVID-19 may develop autoantibodies that target proteins involved in the humoral or cellular response, resulting in decreased levels of B cells or T cells^49^. Similarities to Kawasaki’s disease have led some to propose that multisystem inflammatory syndrome in children (MIS-C) and in adults (MIS-A), rare complications of SARS-CoV-2 infection, may result from aberrant T- or B-cell responses to the virus^35,36^. A subset of patients with COVID-19 also present with cardiomyopathy, viral myocarditis or one of a spectrum of syndromic features associated with “long COVID,” all of which have been linked to immune dysfunction^39^. Comprehensive, high-throughput methods of interrogating the cellular immune response in these conditions can provide important clinical insights.

We acknowledge several study limitations, including small samples sizes in some cohorts tested (<15 days post-symptom onset), very limited data from pediatric cohorts (<18yo), and the lack of availability of other seasonal human coronavirus (HCoV) samples for cross-reactivity testing. For the latter, we made extensive efforts to locate retrospective samples for subjects with common respiratory infections but were unsuccessful as blood is not commonly drawn in the clinical diagnosis or treatment of these respiratory viruses. Importantly, there is a high reported prevalence of antibodies against each of the four HCoVs, with greater than 98% of individuals displaying antibodies against 3 of the 4 common strains^50^. Therefore, a significant number of our controls would be expected to have immune responses against HCoVs, adding confidence to the specificity of our TCR signal.

Diagnostic, therapeutic, and vaccine development for COVID-19 have proceeded at unprecedented speed and scale. T-Detect COVID is the first TCR sequencing-based assay for interrogation of the cellular immune response in SARS-CoV-2, which demonstrates ≥95% positive agreement in identifying prior exposure/infection with ∼100% negative agreement and equivalent or higher performance than commercial EUA serologic testing. As such, it can provide critical insights into disease pathogenesis, severity, recovery, and protection. Future studies will help establish the merits of this approach for immunology research, vaccine/drug development, and public health/surveillance strategies.

## Data Availability

Data requests may be submitted for consideration to: https://www.adaptivebiotech.com/medical-information-request/

## ACKNOWLEDGEMENTS

We thank Kristin MacIntosh (Adaptive) and Melanie Styers (BluPrint Oncology Concepts) for editorial support.

## Supplementary Material

### Detailed Study Protocol Information

All samples were collected pursuant to an Institutional Review Board (IRB)-approved clinical study protocol, “ImmuneSense™ COVID-19 Study” (PRO-00781/ADAP- 007/WIRB#20202820/NCT04583982.) Residual samples collected under prospective study protocols obtained informed consent from participants under a separate protocol: “ImmuneRACE” (ADAP-006/WIRB# 20200625/NCT04494893). All other samples from cohorts described were collected as clinical remnant samples.

### Detailed Methods

#### Description of cohorts used for secondary analyses

The ImmuneRACE study is a prospective, multi-cohort, exploratory study of participants exposed to, infected with, or recovering from COVID-19 (NCT04494893). Participants from across the United States were consented and enrolled via a virtual study design, with cohorting based on participant-reported clinical history following the completion of both a screening survey and study questionnaire. Whole blood, serum, and a nasopharyngeal or oropharyngeal swab were collected from participants by trained mobile phlebotomists. Participants with a confirmed SARS-CoV-2 test were included as residual, retrospective samples in the CV study.

The ImmuneSense™ COVID-19 Study’s prospective study arm enrolled individuals with symptoms suggestive of COVID-19 who were being tested for SARS-CoV-2 at two drive-thru testing sites in New Jersey. Whole blood, serum, and a nasopharyngeal swab were collected from participants at study sites. An electronic questionnaire was administered by study staff. Individuals testing positive via Abbot’s RT PCR were included in the secondary PPA analysis. Individuals testing negative for SARS-CoV-2 using RT-PCR EUA, BioFire RP V2.1, and EUA antibody tests were included in the NPA analysis.

#### Clinical Specimens

From all sources, whole blood samples were collected in EDTA tubes, frozen, and shipped to Adaptive for immunosequencing. When paired serum samples were collected, they were tested using two different EUA antibody assays: 1) Elecsys® AntiSARS-CoV-2; Roche: qualitative detection of high affinity antibodies to SARS-CoV-2 including all isotypes, but preferentially detects IgG antibodies (https://www.labcorp.com/tests/164068/sars-cov-2-antibodies); and 2) SARS-CoV-2 Antibody, IgG; LabCorp: qualitative detection of IgG antibodies to SARSCoV-2 (https://www.labcorp.com/tests/164055/sars-cov-2-antibody-igg).

**Supporting Table 1:**
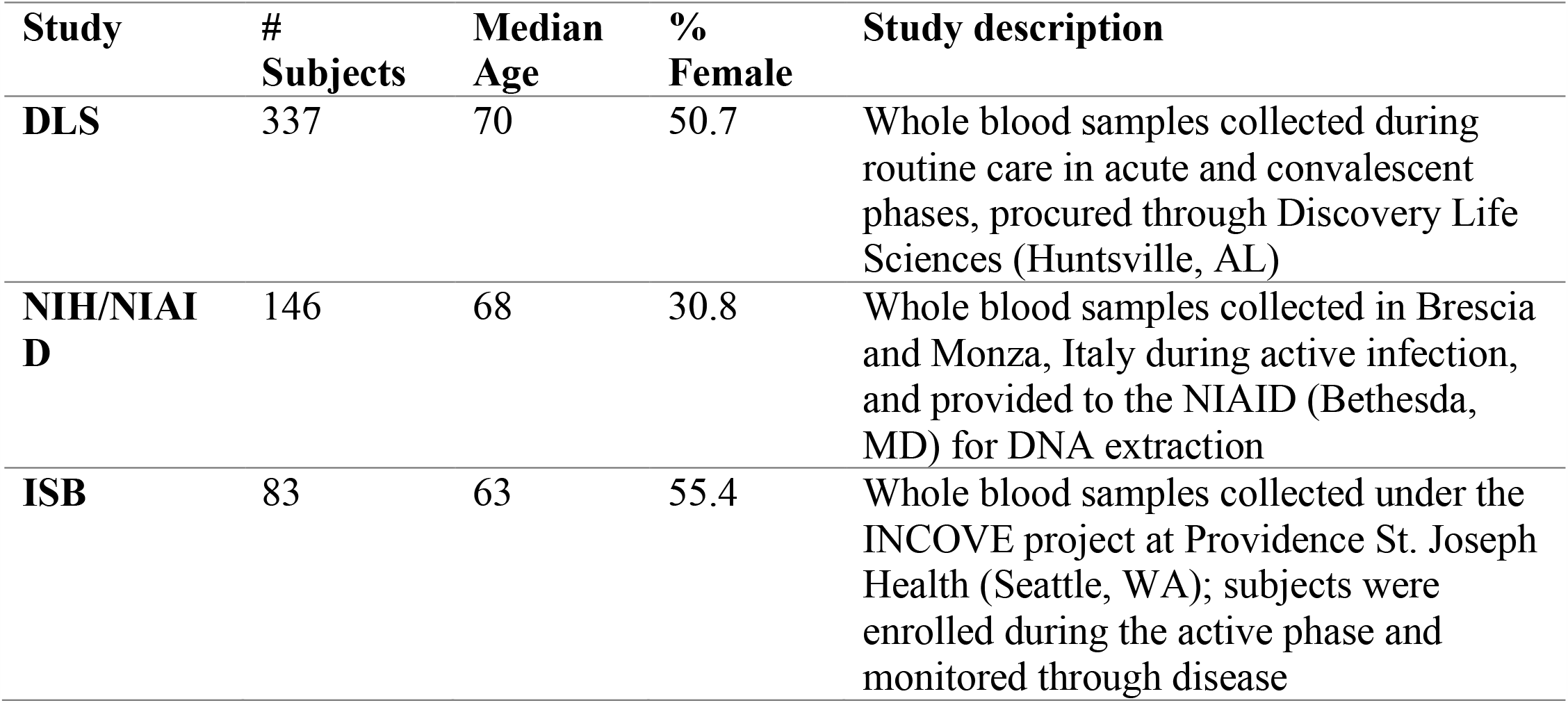
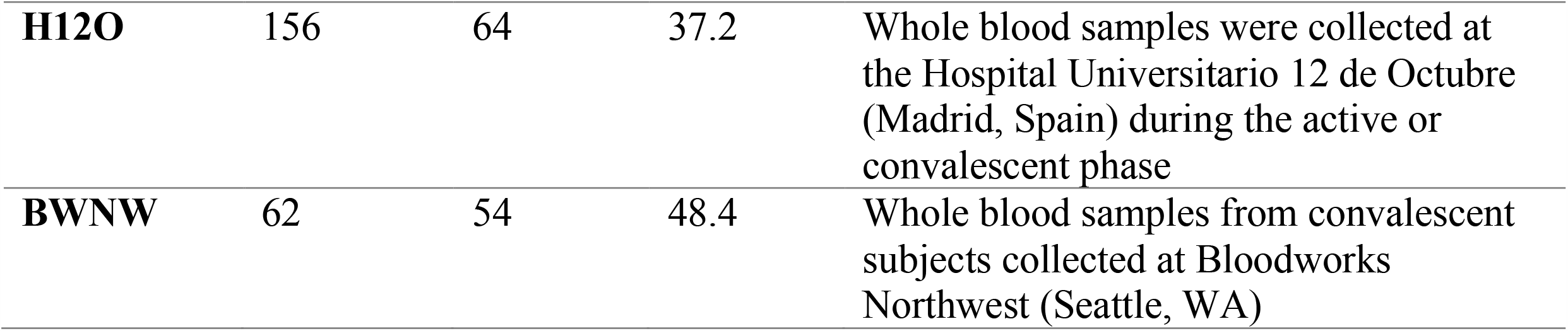
Summary of COVID cohorts used for training of the T-Detect™ COVID classifier:

## Notes

### Competing Interest Statement

SCD declares employment and equity ownership with Adaptive Biotechnologies and employment with Stanford University School of Medicine.
TM and LB declare leadership, employment, and equity ownership with Adaptive Biotechnologies.
All other authors declare employment and equity ownership with Adaptive Biotechnologies.

### Clinical Trial

NCT04583982, NCT04494893

### Funding Statement

Funding was provided by Adaptive Biotechnologies.

### Author Declarations

All samples were collected pursuant to an Institutional Review Board (IRB)-approved clinical study protocol, ImmuneSense COVID-19 Study (PRO-00781/ADAP-007/WIRB#20202820/NCT04583982.) Residual samples collected under prospective study protocols obtained informed consent from participants under a separate protocol: ImmuneRACE (ADAP-006/WIRB# 20200625/NCT04494893). All other samples from cohorts described were collected as clinical remnant samples.

## REFERENCES

1. Emerson, R. O. et al.. Immunosequencing identifies signatures of cytomegalovirus exposure history and HLA-mediated effects on the T cell repertoire. Nat. Genet. 49, 659–665 (2017).

2. Snyder M, T. et al. Magnitude and dynamics of the T-cell response to SARS-CoV-2 infection at both individual and population levels. medRxiv 1–33 (2020) doi:10.1101/2020.07.31.20165647.

3. Johns_Hopkins_University_of_Medicine. Johns Hopkins University of Medicine Coronavirus Resource Center. https://coronavirus.jhu.edu/map.html.

4. Barret, J. R. et al.. Phase 1/2 trial of SARS-CoV-2 vaccine ChAdOx1 nCoV-19 with a booster dose induces multifunctional antibody responses. Nat. Med. (2020) doi:10.1038/s41591-020-01179-4.

5. Huang, A. T. et al.. A systematic review of antibody mediated immunity to coronaviruses: kinetics, correlates of protection, and association with severity. Nat. Commun. 11, 1–16 (2020).

6. Ovsyannikova, I. G., Haralambieva, I. H., Crooke, S. N., Poland, G. A. & Kennedy, R. B. The role of host genetics in the immune response to SARS-CoV-2 and COVID-19 susceptibility and severity. Immunol. Rev. 296, 205–219 (2020).

7. Alter, G. & Seder, R. The power of antibody-based surveillance. N. Engl. J. Med. 383, 1780–1782 (2020).

8. Herroelen, P. H., Martens, G. A., De Smet, D., Swaerts, K. & Decavele, A. S. Humoral immune response to SARS-CoV-2. Am. J. Clin. Pathol. 154, 610–619 (2020).

9. Jacofsky, D., Jacofsky, E. M. & Jacofsky, M. Understanding antibody testing for COVID-19. J. Arthroplasty 35, S74–S81 (2020).

10. Gudbjartsson, D. F. et al.. Humoral immune response to SARS-CoV-2 in Iceland. N. Engl. J. Med. 383, 1724–1734 (2020).

11. Long, Q. X. et al.. Clinical and immunological assessment of asymptomatic SARS-CoV-2 infections. Nat. Med. 26, 1200–1204 (2020).

12. Milani, G. P. et al.. Serological follow-up of SARS-CoV-2 asymptomatic subjects. Sci.Rep. 10, 1–7 (2020).

13. Ward, H. et al.. Declining prevalence of antibody positivity to SARS-CoV-2: a community study of 365,000 adults. medRxiv (2020) doi:10.1101/2020.10.26.20219725v1.

14. Seow, J. et al.. Longitudinal observation and decline of neutralizing antibody responses in the three months following SARS-CoV-2 infection in humans. Nat. Microbiol. 5, 1598–1607 (2020).

15. Deeks, J. J. et al.. Antibody tests for identification of current and past infection with SARS-CoV-2. Cochrane Database Syst. Rev. 2020, (2020).

16. Tzouvelekis, A., Karampitsakos, T., Krompa, A., Markozannes, E. & Bouros, D. False Positive COVID-19 Antibody Test in a Case of Granulomatosis With Polyangiitis. Front. Med. 7, 1–4 (2020).

17. To, K. K. et al.. False-positive SARS-CoV-2 serology in 3 children with Kawasaki disease. Diagn. Microbiol. Infect. Dis. 98, 115141 (2020).

18. Whitman, J. D. et al.. Evaluation of SARS-CoV-2 serology assays reveals a range of test performance. Nat. Biotechnol. 38, 1174–1183 (2020).

19. Grifoni, A. et al.. Targets of T Cell Responses to SARS-CoV-2 Coronavirus in Humans with COVID-19 Disease and Unexposed Individuals. Cell 181, 1489-1501.e15 (2020).

20. Sekine, T. et al.. Robust T cell immunity in convalescent individuals with asymptomatic or mild COVID-19. Cell 183, 158–168 (2020).

21. Le Bert, N. et al.. SARS-CoV-2-specific T cell immunity in cases of COVID-19 and SARS, and uninfected controls. Nature 584, 457–462 (2020).

22. West, R., Kobokovich, A., Connell, N. & Gronvall, G. K. COVID-19 Antibody Tests: A Valuable Public Health Tool with Limited Relevance to Individuals. Trends Microbiol. xx, 1–10 (2020).

23. Tillett, R. L. et al.. Genomic evidence for reinfection with SARS-CoV-2: a case study. Lancet Infect. Dis. 21, 52–58 (2020).

24. Cohen, J. I. & Burbelo, P. D. Reinfection with SARS-CoV-2: Implications for Vaccines. Clin. Infect. Dis. (2020) doi:10.1093/cid/ciaa1866.

25. Staines, H. M. et al.. IgG Seroconversion and Pathophysiology in Severe Acute Respiratory Syndrome Coronavirus 2 Infection. Emerg. Infect. Dis. 27, 85–91 (2021).

26. Pollán, M. et al.. Prevalence of SARS-CoV-2 in Spain (ENE-COVID): a nationwide, population-based seroepidemiological study. Lancet 396, 535–544 (2020).

27. Del Valle, D. M. et al.. An inflammatory cytokine signature predicts COVID-19 severity and survival. Nat. Med. 26, 1636–1643 (2020).

28. Peng, Y. et al.. Broad and strong memory CD4+and CD8+T cells induced by SARS-CoV- 2 in UK convalescent COVID-19 patients. Nat. Immunol. 21, 1336–1345 (2020).

29. Rydyznski Moderbacher, C. et al. Antigen-specific adaptive immunity to SARS-CoV-2 in acute COVID-19 and associations with age and disease severity. Cell 183, 996-1012.e19 (2020).

30. Wyllie, D. et al.. SAR-CoV-2 responsive T cell numbers are associated with protection from COVID-19:A prospective cohort study in keyworkerd. medRxiv (2020) doi:10.1101/2020.11.02.20222778.

31. Funk, C. D., Laferrière, C. & Ardakani, A. A snapshot of the global race for vaccines targeting SARS-CoV-2 and the COVID-19 pandemic. Front. Pharmacol. 11, 1–17 (2020).

32. Schulien, I. et al.. Characterization of pre-existing and induced SARS-CoV-2-specific CD8+ T cells. Nat. Med. (2020) doi:10.1038/s41591-020-01143-2.

33. Zuo, J. et al.. Robust SARS-CoV-2-specific T-cell immunity is maintained at 6months following primary infection. bioRvix (2020) doi:10.1101/2020.11.01.362319.

34. Gallais, F. et al.. Intrafamilial Exposure to SARS-CoV-2 Induces Cellular Immune Response without Seroconversion. medRxiv 1–15 (2020) doi:10.1101/2020.06.21.20132449.

35. Levin, M. Childhood multisystem inflammatory syndrome–A new challenge in the pandemic. N. Engl. J. Med. 383, 393–395 (2020).

36. Weatherhead, J. E., Clark, E., Vogel, T. P., Atmar, R. L. & Kulkarni, P. A. Inflammatory syndromes associated with SARS-CoV-2 infection: dysregulation of the immune response across the age spectrum. J. Clin. Invest. 130, 6194–6197 (2020).

37. Varga, Z. et al.. Endothelial cell infection and endotheliitis in COVID-19. Lancet 395, 1417–1418 (2020).

38. Siripanthong, B. et al.. Recognizing COVID-19–related myocarditis: The possible pathophysiology and proposed guideline for diagnosis and management. Hear. Rhythm 17, 1463–71 (2020).

39. Marshall, M. The lasting misery of coronavirus long-haulers. Nature 585, 339–341 (2020).

40. DeFrancesco, L. Whither COVID-19 vaccines? Nat. Biotechnol. 38, 1132–1145 (2020).

41. Korber, B. et al.. Tracking Changes in SARS-CoV-2 Spike: Evidence that D614G Increases Infectivity of the COVID-19 Virus. Cell 182, 812-827.e19 (2020).

42. Gittelman, R. M. et al.. Diagnosis and tracking of past SARS-CoV-2 Infection in a large study of Vo’, Italy through T-cell receptor sequencing. medRxiv 2–12 (2020) doi:10.1101/2020/11/09.20228023.

43. In Vitro Diagnostics EUAs (FDA website). https://www.fda.gov/medical-devices/coronavirus-disease-2019-covid-19-emergency-use-authorizations-medical-devices/vitro-diagnostics-euas.

44. Zuo, J. et al.. Robust SARS-CoV-2-specific T-cell immunity is maintained at 6 months following primary infection. bioRxiv 2020.11.01.362319 (2020) doi:10.1101/2020.11.01.362319.

45. Anderson, E. J. et al.. Safety and immunogenicity of SARS-CoV-2 mRNA-1273 vaccine in older adults. N. Engl. J. Med. 2427–2438 (2020) doi:10.1056/nejmoa2028436.

46. Widge, A. et al.. Durability of responses after SARS-CoV-2 mRNA-1273 vaccination. N. Engl. J. Med. NEJMc20321, Epub ahead of print (2020).

47. Ewer, K. et al.. T cell and antibody responses induced by a single dose of ChAdOx1 nCoV-19 (AZD1222) vaccine in a Phase 1/2 clinical trial. Nat. Med. (2020) doi:10.1038/s41591-020-01194-5.

48. Lambert, P.-H. et al.. Consensus summary report for CEPI/BC March 12–13, 2020 meeting: Assessment of risk of disease enhancement with COVID-19 vaccines. Vaccine In press, (2020).

49. Wang, E. Y. et al.. Diverse functional autoantibodies in patients with COVID-19. medRxiv (2020) doi:10.1101/2020.12.10.20247205;

50. Gorse, G. J., Patel, G. B., Vitale, J. N. & O’Connor, T. Z. Prevalence of antibodies to four human coronaviruses is lower in nasal secretions than in serum. Clin. Vaccine Immunol. 17, 1875–1880 (2010).

